# Improving prediction of amyloid deposition in Mild Cognitive Impairment with a timed motor task

**DOI:** 10.1101/2021.06.16.21259056

**Authors:** Sydney Y. Schaefer, Kevin Duff, Andrew Hooyman, John M. Hoffman

## Abstract

Cortical amyloid deposition is one of the hallmark biomarkers of Alzheimer’s disease. However, given how cost- and time-intensive amyloid imaging can be, there is a continued need for a low-cost, non-invasive, and accessible enrichment strategy to pre-screen individuals for their likelihood of amyloid prior to imaging. Previous work supports the use of coordinated limb movement as a potential screening tool, even after controlling for cognitive and daily function. Thirty-six patients diagnosed with amnestic Mild Cognitive Impairment over the age of 65 underwent ^18^F-Flutemetamol amyloid-positron emission tomography imaging, then completed a timed motor task involving upper limb coordination. This task takes ∼5 minutes to administer and score. Multivariate linear regression and Receiver Operator Characteristic analyses showed that including motor task performance improved model prediction of amyloid burden. Results support the rationale for including functional upper extremity motor assessment as a cost- and time-effective means to screen participants for amyloid deposition.

## INTRODUCTION

Cortical amyloid deposition is one of the hallmark biomarkers of Alzheimer’s disease (AD) and its progression ^1,2^. Thus, numerous large-scale clinical trials in preclinical AD have focused on therapies aimed at clearing beta-amyloid neuritic plaques to slow disease progression. However, recruiting and enrolling asymptomatic individuals who are amyloid positive is time-consuming, since only ∼30% of cognitively-intact individuals have elevated levels of amyloid ^3^. This means that two out of every three individuals who undergo amyloid positron emission tomography (PET) as part of the screening process for clinical trial recruitment will not be eligible for enrollment. Furthermore, amyloid imaging is expensive, exposes individuals to radiation, and can only be completed select sites with the necessary technology and expertise. Thus, there is a need for a low-cost, non-invasive, and accessible way to pre-screen asymptomatic individuals for their likelihood of β-amyloid neuritic plaque density prior to PET imaging.

Although complex movements involving multilimb coordination have been associated with disease severity ^4-6^, recent work has also demonstrated that such movement may be sensitive to disease *progression* ^7^ when assessed with a timed motor task. To minimize cost and assessment time and improve portability, we developed an upper extremity motor task that *i)* does not require any hardware or software; *ii)* can differentiate between cognitively intact and cognitively impaired individuals ^8^ better than other simple motor tasks (i.e., grip strength, see ^9^); and *iii)* is feasible for amnestic Mild Cognitive Impairment (MCI) cohorts ^7,10^. This is in contrast to other assessments of complex movement that require demanding technology (e.g., movement sensors ^6^, motion capture technology ^5^, electromyography ^4^, or transcranial magnetic stimulation ^11^) or do not show strong prognostic effects at baseline (e.g., ^12^). Given the relative advantages of this timed motor task and its prediction of functional decline in MCI, we hypothesized that task performance would be related to the extent of amyloid plaque deposition, and would improve the classification of amyloid positivity in individuals with amnestic MCI, above and beyond baseline cognitive and activities of daily living.

## MATERIALS & METHODS

### Participants

Thirty-six participants with amnestic MCI from a larger clinical trial sample (ClinicalTrials.gov Identifier: NCT02301546; currently active, not recruiting) participated (mean±SD age = 73.25±5.5 years; 13 females; 16.81±3.0 years of education; 97% white). Inclusion criteria were 65 years old or older, had a collateral source available to answer questions about thinking abilities and daily activities, had access and the ability to use a computer and the internet, spoke English, and demonstrated that they had single- or multi-domain amnestic MCI. MCI was categorized as: 1) concern of a change in cognition from the participants or a knowledgeable informant, 2) impairment in memory (and other cognitive domains), with at least one cognitive test score in a domain being 1.5 standard deviations below an estimate of premorbid intellect, and 3) independence of daily functioning ^13^. Exclusion criteria were history of major neurological (e.g., stroke, Parkinson’s disease) or psychiatric illnesses (e.g., schizophrenia, bipolar disorder) or substance abuse, current major depression (>7 on the 15-item Geriatric Depression Scale), or cognitive impairment suggestive of dementia. This study was approved by the University of Utah Institutional Review Board, in accordance with the World Medical Association Declaration of Helsinki. All participants provided informed consent as self or by proxy prior to enrollment.

### Timed motor task

A full visual description of the timed motor task can be viewed on Open Science Framework (https://osf.io/phs57/wiki/Functional_reaching_task/), and its methods have been published previously ^7-10^. To summarize, participants use a standard plastic spoon to acquire two raw kidney beans at a time from a central cup (all cups 9.5cm diameter and 5.8cm deep) to one of three distal cups arranged at a radius of 16 cm at -40°, 0°, and 40° relative to the central cup. All cups were the same size. Participants were tested using their nondominant hand, and started by moving to the cup ipsilateral (same side) of the hand used. They then returned to the central cup to acquire two more beans at a time to transport to the middle cup, then the contralateral cup, and then repeated this sequence four more times for a total of 15 out-and-back movements. Task performance was measured as trial time (in seconds), i.e., how long it took to complete 15 movements, such that lower values indicate better performance. Movement errors, such as dropping beans mid-reach, were recorded; however, only 1 error (0.1% of all reaches) was made in this dataset. Participants first completed 3 trials for practice and task familiarization.

### Amyloid-PET imaging

Participants received ^18^F-Flutemetamol imaging as described previously ^14. 18^F-Flutemetamol was produced under PET cGMP standards and the studies were conducted under an approved Federal Drug Administration Investigational New Drug application. Imaging was performed 90 minutes after the injection of 185 mBq (5 mCi) of ^18^F-Flutemetamol. Emission imaging time was approximately 20 minutes. A GE Discovery PET/CT 710 (GE Healthcare) was used in this study. This PET/CT scanner has a full width at half-maximum spatial resolution of 5.0 mm and excellent performance characteristics ^15,16. 18^F-Flutemetamol uptake was analyzed using a regional semi-quantitative technique ^17,18^. In this technique, semi-quantitative regional (prefrontal, anterior cingulate, precuneus/posterior cingulate, parietal, mesial temporal, lateral temporal, occipital, sensorimotor, cerebellar grey matter, and whole cerebellum) regional standardized uptake value ratios (SUVR) were generated automatically and normalized to the pons. Based on the regional values a composite standardized uptake value ratio (composite SUVR) of the cerebral cortex was generated automatically and normalized to the pons using the CortexID Suite software ^19^. This software uses a threshold *z* score of 2.0 to indicate abnormally increased regional amyloid burden that corresponds to a composite SUVR of 0.59 when normalized to the pons, providing a 99.4% concordance with visual assessment ^17^. For ^18^F-Flutemetamol amyloid imaging, there is no specific age-related “normal” level of binding in the CortexID Suite database to assess age-matched normality. Thus, the study images were compared to the intrinsic software database control group as a whole to calculate the z-scores compared to clinically negative amyloid scans.

### Measures of cognitive and daily functioning

As part of the clinical trial, participants underwent extensive neuropsychological assessment at baseline; however, only the Delayed Memory Index from the Repeatable Battery for the Assessment of Neuropsychological Status (RBANS, ^20^) was examined here. All subtests were administered and scored as defined in the manual, and normative data from RBANS manual was used to calculate the Index score, which are presented as age-corrected standard score (M = 100, SD = 15) with higher scores indicating better cognition. Mean±SD RBANS Delayed Memory Index scores for this sample were 74.42±21.01, consistent with their diagnosis. Baseline activities of daily living (ADL) function was measured using the self-report portion of the 18-item Alzheimer’s Disease Cooperative Study-Activities of Daily Living scale adapted for MCI (ADCS-ADL-MCI) ^21^. Possible scores on this scale range from 0 to 57, with higher scores indicating better daily functioning. Mean±SD ADCS-ADL-MCI scores were 46.08±3.82, again consistent with their diagnosis.

### Statistical analysis

Multivariate linear regression was conducted to predict ^18^F-Flutemetamol pons normalized composite SUVRs using participants’ motor task performance (i.e., trial time) as a predictor while controlling for age, gender, years of education, RBANS Delayed Memory Index score, and ADCS-ADL-MCI-18 score. Assumptions for regression were inspected visually using Q-Q plots and all analysis were performed in R (v3.5.1). Statistical models with and without motor task performance as a dependent variable were compared by analysis of variance to determine if the contribution of motor task performance to prediction accuracy was statistically significant.

To test whether motor task performance improved amyloid positivity classification (Aβ+ or Aβ-), we first developed a null model using best practices of model selection ^22^ that included age, sex, education, RBANS Delayed Memory Index score, and ADCS-ADL-MCI-18 score. A generalized linear model was selected since amyloid positivity follows a binomial distribution. We then generated a motor task model that included the null model plus the motor task variable. Akaike information criteria (AIC) and analysis of variance (ANOVA) using a Chi-squared distribution were used to test for model superiority (null vs. task). This determined if including motor task performance as a variable improved prediction accuracy of amyloid classification without added model complexity. An AIC difference of >3 between the null and task model would indicate improved data fit by the task model. Receiver operator characteristics (ROC) and precision recall curves were also generated to assess model specificity, sensitivity, precision and recall with and without motor task performance.

## RESULTS

No adverse events were reported during the injection, uptake time, or imaging studies with the investigational imaging agent ^18^F-Flutemetamol. Mean composite of SUVRs normalized to the pons was 0.68 (SD=0.18, range=0.41 – 0.97). Mean motor task performance was 63.88 seconds (SD=15.66, range=39.81 – 121.75). For reference, cognitively-intact older adults tend to be faster (M=58.50 seconds, data from ^8,23^).

Regression analyses revealed that motor task performance was a significant predictor of composite SUVR (β = .004; 95% CI = [.0004, .008]; p = .03), even when controlling for age (p = .17), gender (p = .1), years of education (β = .03; 95% CI = [.013, .05]; p = .002), RBANS Delayed Memory Index score (p = .34), and ADCS-ADL-MCI score (p = .25). The full model yielded an adjusted R^2^ = .25 (F(6,29) = 3.11; p = .022). Comparison of regression models with and without motor task performance (R^2^ =.15; p = 0.08) through analysis of variance demonstrated that the inclusion of motor task performance significantly improved prediction (p = .03) of composite SUVR by over 65%.

Based on established thresholds, 26 of the 36 participants (72%) were classified as amyloid-positive. The best generalized linear model of the covariate data, i.e. the null model, included age, sex, education, RBANS Delayed Memory Index score, and ADCS-ADL-MCI score (AIC = 44.1) as predictors of amyloid positivity classification. Adding motor task performance to the null model improved model accuracy (AIC = 41.4). ANOVA confirmed that the motor task model was more accurate than the null model (p = .03) in predicting amyloid classification.

ROC showed that the motor task model had a specificity of 60% (6/10 prediction accuracy of Aβ-), and a sensitivity of 88% (23/26 prediction accuracy of Aβ+) with an overall accuracy of 75% compared to the null model, which had a specificity of 50% (5/10 prediction accuracy of Aβ-) and a sensitivity of 93% (24/26 prediction accuracy of Aβ+) with an overall accuracy of 80%. Overall, the motor task model had an AUC of 90%, compared to the null model AUC of 84% (Fig. 1A).

**Figure 1.**
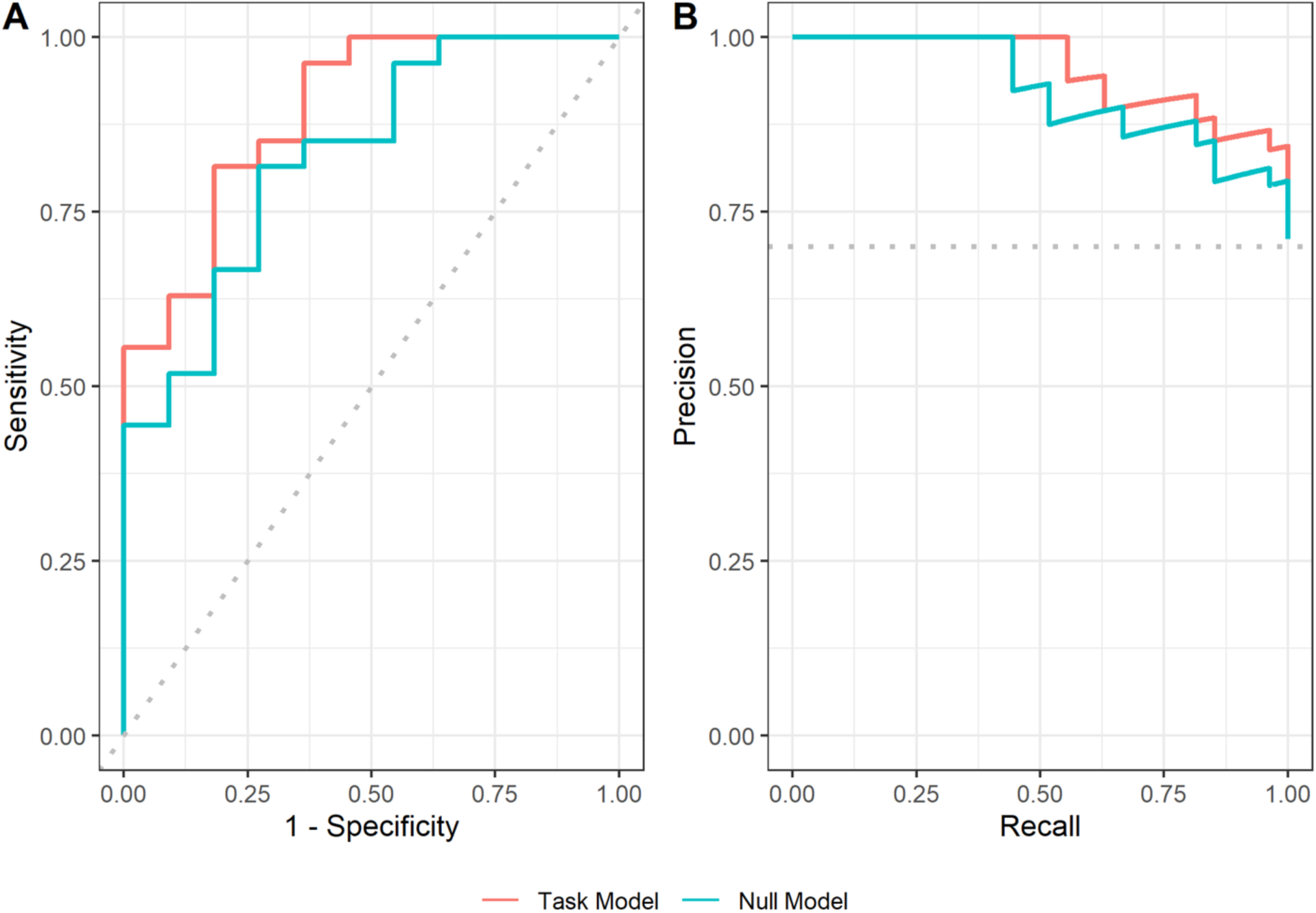
(A) Receiver Operator Characteristic and (B) Precision Recall curves for the null (blue) and motor task (orange) models for predicting the probability of amyloid positivity.

Given that the majority of participants were classified as amyloid positive, precision recall curves (PRC) were also generated for each model ^24^. Briefly, a precision recall curve determines the trade-off of a model between its true-positive rate and its positive prediction rate by varying the ratio between positive and negative cases and assessing the predictive skill of the model throughout. This can be an especially important metric when evaluating samples with a disproportionate number of positive or negative cases ^24^. Here, the area under the PRC of the motor task model was 96% compared to that of the null model, which was 93% (Fig. 1B). This further demonstrates that advantage of including motor task performance for predicting amyloid-positive cases even when the ratio between positive and negative cases may be skewed, such as in preventative clinical trials where the number of amyloid-negative cases is much higher (e.g., ^25^).

To determine an optimal cut-off of motor task performance to predict amyloid-positive cases, a permutation test was run that varied motor task cut-off threshold across the range of performance times observed in this sample, followed by a calculation of the resulting odds ratio for amyloid positivity. The cut-off value with the highest odds ratio was determined to be the optimal threshold, which was a task performance of 68 seconds with an odds ratio of 4.76.

## DISCUSSION

The purpose of this brief report was to test whether performance on a timed motor task was related to the extent of amyloid plaque deposition in individuals with amnestic MCI, and would improve the classification of amyloid positivity. Results showed that even after controlling for age, gender, education, delayed memory, and ADL function, motor task performance was still a significant predictor of composite SUVR, with worse task performance being associated with more amyloid deposition. Furthermore, adding motor task performance as a predictor variable improved amyloid positivity classification, being able to better identify individuals with elevated amyloid than with just age, gender, education, delayed memory, and ADL function. Overall, these findings support the rationale for including functional upper extremity motor assessment as a means to better screen participants for clinical trial recruitment that requires elevated amyloid for enrollment (e.g., Anti-Amyloid Treatment in Asymptomatic Alzheimer’s Disease [A4]).

Although several complex upper extremity motor tasks have been shown to be sensitive to disease severity ^4,6^, this is among the first to show a relationship with disease biomarkers, above and beyond other measures such as memory or ADL function. While this study does not provide a clear mechanism of this relationship, it possible that unimanual motor performance may be sensitive to amyloid deposition patterns in sensory-motor areas specifically ^26^, which may track with global composite measures. It is also likely that this task, more so than grip dynamometry or finger tapping that do not have a strong visuospatial demand, recruits relevant neural structures (e.g., hippocampus) that are particularly susceptible to early stages of dementia ^27,28^. Future research is needed, however, to further explore the underlying mechanism between complex motor tasks and both global and regional amyloid deposition.

It is acknowledged that screening for amyloid deposition is already a time- and cost-intensive process, particularly in mild cases or those who are asymptomatic. Efforts to identify Aβ+ individuals have been enriched by additional biomarkers, genetic testing, and extensive neuropsychological evaluation, which also take time and/or money, and are still not always sensitive and specific to amyloid or disease progression. We therefore highlight the fact that the motor task used in this study takes <5 minutes to administer and costs less than $10 to fabricate from household items, thereby potentially improving the likelihood of identifying individuals with amyloid accumulation with virtually no additional time or cost. It is also extremely portable, with data collection easily available in clinics and the community. In fact, using these time and cost parameters as inputs into the Biomarker Prognostic Enrichment Tool (BioPET) ^29^, along with published rates of amyloid positivity in cognitively-intact adults ^3^, it is estimated (with a power of 0.9) that just by pre-screening individuals with the timed motor task could reduce the total cost for amyloid scanning by ∼36%. For example, in a preventative AD clinical trial that attempts to recruit 1,000 amyloid-positive subjects, this 36% could reflect millions of dollars in savings (as well as countless hours for the study personnel and patients and their families). Furthermore, the task’s extremely low price and rapid testing time compared to amyloid-PET still outweigh the estimated 1.5x increase in total individuals screened, thereby streamlining and improving the efficiency of clinical trial recruitment through additional enrichment strategies.

We acknowledge the high education levels and lack of racial/ethnic diversity within the relatively small sample, which warrant future research in larger and more diverse cohorts to better estimate the potential of motor behavior as an affordable enrichment strategy for AD clinical trials. We also acknowledge that the relative strength of the timed motor task as a predictor of amyloid was not directly compared to other existing motor tasks (e.g., grip dynamometry, 10 Meter Walk Test), but we have previously shown that the motor task presented here is likely more sensitive to disease severity than other motor assessments ^9^ (see also ^30^). As such, motor assessments have promise as cost-effective and non-invasive screening tools that would allow for enriching samples in clinical trials in AD.

## Supporting information

STROBE checklist

## Data Availability

Data are not publicly available.

## ACKNOWLEDGEMENTS

The ADCS-ADL-MCI survey was used with permission from the NIA Alzheimer’s Disease Cooperative Study (NIA Grant AG10483).

